# At what geographic scales does agricultural alienation amplify foodborne disease outbreaks? A statistical test for 25 U.S. states, 1970-2000

**DOI:** 10.1101/2019.12.13.19014910

**Authors:** Kenichi W. Okamoto, Alex Liebman, Robert G. Wallace

## Abstract

The modern economy is driving multiple environmental and social crises across the globe. Capital accumulation externalizes the myriad damage associated with commodity production to ecosystems, labor, public health, and governments across jurisdictions. A growing literature shows multinational agriculture, a major sector of the economy, plays a fundamental role in disrupting the ecological cycles upon which communities across the globe depend. We report here one of the first statistical tests of such an ecosocial rift. We used geometric morphometrics to characterize a parameter space in agricultural alienation across nature, human welfare, and industrial appropriation for 25 U.S. states, at five-year increments between 1970 and 2000. The first two relative warps of the analysis reproduce the long-documented shift in and out of the 1980s farming crisis. We found the crisis left the U.S. food system in a new configuration, with commodity crops replacing cropland pasture, greater farm debt load, overcapitalized inputs, and a relative decline in on-farm wages. To determine if such a shift had an epidemiological impact, we tested whether salmonellosis and shigellosis—two foodborne pathogens for which national incidence data across the study period were available—regressed against historical changes in this alienation space across geographic scale. The second relative warp for shigellosis showed a multivariate relationship with the alienation space, but significance failed to withstand a Bonferroni correction. The partial warps and Procrustes residuals of temporal trends in the alienation space likewise exhibited limited predictive capacity for outbreak size. In part these results may reflect sampling artifacts. Data availability limits both the number of years sampled and the variety of diseases tested. Nevertheless, our analyses demonstrate that metabolic rifts associated with specific modes of production can be rigorously investigated.

## Introduction

The present environmental crisis is unprecedented in its global impact upon humanity (Intergovernmental Panel on Climate Change 2014, 2018, Kolbert 2014, Dawson 2016, Wallace-Wells 2019). Climate change, deforestation, ocean acidification, pollution, nitrate and phosphate loading, freshwater depletion, and disruptions in thermohaline circulation have either surged across ecological tipping points or are rapidly approaching them (Rockstrom et al. 2009, 2009b, Barnosky et al. 2012, RG Wallace and Kock 2012, Hughes et al. 2013, Allen et al. 2015, Drijhout et al. 2015, Rocha et al. 2015, Diamond et al. 2015, Dakos et al. 2019). The resulting weather shifts, habitat destruction, biodiversity loss, ecosystem dysfunction, disease emergence, resource depletion, eutrophication, soil degradation, fisheries collapse, and environmental toxicity have struck together in short order by geological and even historical standards (Mac Nally et al. 2009, Carpenter et al. 2011, Choat et al. 2012, Enering et al. 2013, Myers et al. 2014, Gauthier et al. 2015, Ripple et al. 2015, Essington et al. 2015, Peres et al. 2016, Perry et al. 2017, Zhou et al. 2018, Anderson et al. 2018, Filippelli and Taylor 2018). Many of the regional biomes upon which human communities across the world depend are now under existential threat.

The danger is unlikely a deterministic consequence of humanity’s putative nature (Malm and Hornborg 2014). Instead, the ongoing crisis has repeatedly been connected to a particular mode of social reproduction (McIntosh et al. 2000, Chakrabarty 2009, Büntgen et al. 2011, Moore 2017). Centers of capital accumulation and their multinational and State organizations have driven unprecedented increases in resource extraction, per capita consumption, and unabsorbed wasteload (Meszaros 1970, O’Hara 2009, Foster et al. 2010, Heede 2014, Stehle and Schulz 2015, Moore 2015, Baer and Singer 2016, Malm 2016, Bergmann and Holmberg 2016, Hamilton et al. 2018).

Although one among many modes of production, the agricultural sector represents a particularly sharp pressure upon the biosphere’s capacity to regenerate dynamic ecosystemic networks (Foley et al. 2005, Ellis and Ramankutty 2008, Alexandratos and Bruinsma 2012, FAO 2013, R Wallace et al. 2016, Ramankutty et al. 2018, Vandermeer et al. 2018, GRAIN and IATP 2018). In focusing its operations almost entirely around turning natural and social resources into commodities, industrial agriculture has decoupled ecological feedback from social metabolism (Mancus 2007, Gunderson 2011, RG Wallace and Kock 2012, Clausen et al. 2015, Sharp 2016, Foster 2016, Schneider 2017, RG Wallace et al. in preparation). The resulting metabolic rifts manifest in destruction across geological, ecological, and public health domains, including global warming, polluted air and waterways, reduced agrobiodiversity, pollinator collapse, new infectious diseases, declines in crop nutrition, globalized morbid obesity and diabetes, farm consolidation and dispossession, widespread antibiotic resistance, farmer suicides, and an attendant opioid epidemic (Potts et al. 2010, Pereira et al. 2012, da Costa et al. 2013, Jones et al. 2013, Malik et al. 2013, Lowder et al. 2016, IPES-Food 2016, Mateo-Sagasta et al. 2017, Bryant and Garnham 2017, Peters et al. 2019, Tessum et al. 2019, Intergovernmental Panel on Climate Change 2019).

The ecosocial alienation that this series of capital-led crises imposed upon humanity, nature, and our capacity to appropriate natural resources has been treated almost entirely in qualitative terms. Attempts to quantify such disjunction include Clement (2009), who used robust regression to evaluate how Texas county demographics within a single year related to municipal solid-waste, finding population per square mile and per capita income significant. By a fixed-effects panel regression, controlling for time-constant features within subject and events factors shared across subjects, McGee and Alverez (2016) tested for biochemical oxygen demand, a marker of water pollution, in organic farming across countries. The two analyses illustrate that one can test for alienation across land-use patterns, human activity, and human welfare over short-to-medium time horizons. Here we offer what is, to our knowledge, one of the first attempts at statistically testing such rifts between human production and nature spanning several decades.

Such a research program is not a trivial enterprise, as much for the questions to be formulated as for its technical difficulties. A disembedded economy entails a complex interplay of social and natural processes (Mészáros 1970, Harvey 1993 [2016], Lewontin 1998, Levins and Lopez 1999, RG Wallace et al. in press). Understanding these causal webs in such a high-dimensional space requires methods that can capture the essential structure of the study system’s complexity while also accounting for its changes. A variety of multivariate methods—life cycle analyses, ecological footprinting, and input-output analyses of industrial ecology—are available to track the effects of capital production and circulation on land and labor alike.

For instance, Bergmann and Holmberg (2016) identified the croplands, pastures, and forests that are part of commodity webs supporting export-oriented development across international markets. Bergmann and Holmberg (2016) further differentiated foreign consumption/accumulation of “direct” agricultural goods; processed agricultural goods; manufactured goods as far afield as electronics and vehicles; and services, including air transport, insurance, and education. Bergmann (2013, 2017) extended these relational geographies to carbon emissions and labor intensity.

Here we apply an approach whose virtue lies in both visualizing these higher-order interactions and statistically testing their impacts upon potential response variables. Geometric morphometrics, originally developed for characterizing the morphologies of individual organisms, has been applied to other parameter spaces, including the kind of geographic data we analyze here (Tobler 1994, RG Wallace 2002, Yabe 2005, O’Sullivan et al. 2017, Bookstein 2018). The approach’s statistics trace how the aggregate of data points—the interrelationships among vertices—shifts over time. We use geometric morphometrics to determine in which directions an agricultural three-dimensional space defined across nature, human welfare, and industry for 25 U.S. states underwent alienation between 1970 and 2000. We also assess whether changes in alienation space had a public health impact. In particular, we test whether national incidences for two foodborne pathogens, salmonellosis and shigellosis, significantly regress with shifts in U.S. agricultural alienation. Our aim was to devise a systematic framework for evaluating at which geographic scales agricultural alienation amplified these outbreaks.

## Methods

### Agricultural alienation parameter space

We first constructed a parameter space defining alienation in agriculture by U.S. state in the period 1970-2000 across three axes: nature, human, and industry (Figure 1). The 25 states were selected by an allometric criterion of at least six million acres of cropland as of 1945.

**Figure 1.**
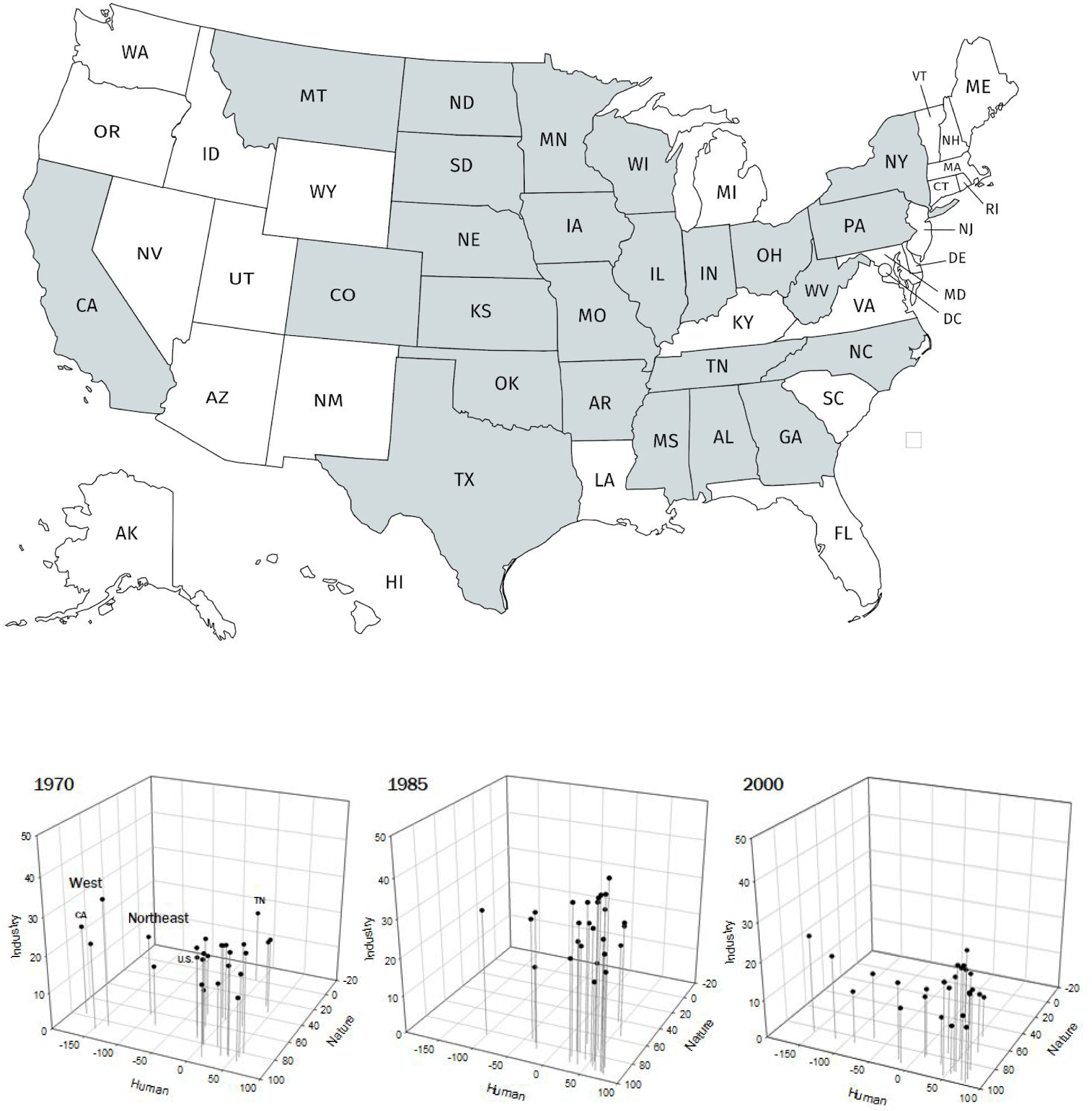
Agriculture alienation across 25 U.S. states, 1970-2000. Study states in grey at top. We tracked the system every five years, but in the interests of concision show only three years here. The West—California, Colorado, and Montana—and the Northeast—New York and Pennsylvania—largely group to themselves, while states of the Upper Midwest, the Corn Belt, the South/Appalachia, and the Southern Plains/Delta, above the U.S. average and on the leading edge of industrial production, group toward the front right corner. A negative nature alienation marks a single case wherein cropland pasture slightly exceeded commodity cropland (Tennessee, 1970), marking the end of an era on the Southern Plains. Negative human alienation, with wages exceeding fertilizer costs, marks no socialistic turn, only that fertilizers had not been applied to the extent to which they came to dominate production by 2015 and/or that human labor still served as an important source of productive power on-site. California and Colorado, for instance, remained in the negative for human alienation as defined here for the entirety of the study period.

*Nature alienation* in reference to agriculture was characterized as the complement percent of a state’s ratio of cropland used for pasture to cropland for crops or pasture-to-cropland (1 - *p/c*). These land use data were sourced from the Economic Research Service (ERS) of the U.S. Department of Agriculture for years 1969 (assigned to 1970), 1974 (1975), 1978 (1980), 1987 (1985), 1992 (1990), 1997 (1995), and 2002 (2000). Negative percents indicate cropland pasture exceeds annual crops. The ERS notes that for these data, cropland includes crops harvested, crop failure, and summer fallow. Estimates prior to 2012 were based on Nickerson et al. (2011), Lubowski et al. (2006), Vesterby and Krupa (2001), Daugherty (1991 and 1995), Frey and Hexem (1985), Frey (1973, 1979, and 1982), Frey et al. (1968), Wooten et al. (1962), Wooten and Anderson (1957), and Wooten (1953). Estimates for 2012, eventually dropped from this analysis, were based on USDA-Economic Research Service calculations using USDA-National Agricultural Statistics Service (2012, 2014a, and 2014b) and USDA-Farm Service Agency (2014). Changes in pasture should capture the state of system diversity, the scale of conversion to industrial fiber and grains, and the metabolic rifts produced in changing livestock diet to grain and removing large animals—sources of manure and promoters of carbon sequestration—from the open landscape (González 2012).

*Human alienation* is represented here by the ratio of farmer wages-to-fertilizer expenditures (1 – *w/f*). This variable offers a proxy for the gap between investment in production and wage labor’s capacity to reproduce itself, an agricultural analog to Piketty’s (2014) capital-to-income ratio. These data were drawn from the Bureau of Economic Analysis of the U.S. Department of Commerce, and exclude off-farm income sources. *Industry alienation* is measured here by farm debt-to-equity. This ratio represents the extent to which farm revenue is available to producers or passed onto paying off mortgages and/or buying corporate livefeed, fertilizer, pesticide, seed, machinery, and other inputs and services that farmers once drew from nature and labor on-site. These data were drawn from the Economic Research Service of the U.S. Department of Agriculture, and, for debt data prior to 1994, the Farmers Home Administration.

Finally we evaluate the relationship between agricultural alienation and disease outbreak, one hypothesized ramification of increasing alienation across the three dimensions. We use a time series in nationwide annual incidence for two foodborne bacterial pathogens, *Salmonella* and *Shigella*, collated from Groseclose et al. (2003).

### Alienation shape projection

#### Consensus configuration

Each year in the alienation parameter space was treated as a separate *specimen*. Each specimen’s shape was characterized by the configuration across the data points or *landmarks*. These points correspond to geographic entities—U.S. states—and they are quantified along the axes of “human,” “nature,” and “industry” represented in Figure 1. The data depicted in the figure were converted into a three-dimensional landmark dataset using the arrayspecs function from the R package geomorph (Adams et al. 2019). We applied a generalized *Procrustes transformation* to this landmark dataset using the gpagen function, also from geomorph. The method aims to iterate through the specimens and remove the effects of contingent differences in rotation, scaling, and translation among the aggregations of data points, ensuring that the resulting variation among years is characterized exclusively by variation in the configuration of data in our projected alienation space.

The gpagen function also generates a *consensus shape*—namely, a configuration that represents the average arrangement of landmarks among specimens in our three-dimensional setting. In anatomical morphometrics, this allows each specimen’s biological shape to be characterized by the extent to which it differs from the consensus shape (Bookstein 1997, Bookstein 2018). In our application, a comparison of the consensus shape with specific years allows us to visualize how the arrangement of data points projected onto our three-dimensional characterization of shifts in multifaceted alienation over time.

#### Partial and relative warps

If our data points or landmarks are envisioned to occupy a Cartesian, three dimensional space, the extent to which each specimen’s or, in this case, year’s shape deviates from the consensus shape can be described by the deformation needed to produce each specimen’s shape from the consensus shape. Axes that highlight key differences among all specimens constitute the principal components, or eigenvectors, of the covariance matrix among samples at increasing scales of nonaffine effect, from the most local among the smallest subsets of landmarks up to the more global across all the landmarks. These *partial warps*, however, are usually still correlated (Rohlf 1998). To capture the variation among our sample years along orthogonal axes, and so reduce data dimensionality, the partial warp scores can be subjected to a singular value decomposition (Bookstein 1991). The resulting *relative warps* are the linear combinations of partial warps, each warp normalized to a length of one across all specimens. Because Procrustes analyses standardize the landmarks, the axes generated in this process can be viewed as major gradients along which the shapes of our specimen-years alone differ.

We calculated the partial warps using the procGPA function provided by the shapes package in R (Dryden 2018). A single partial warp accounted for approximately 35% of shape variability. Only two other partial warps accounted for more than 5% of shape variability. The top five partial warps accounted for approximately 57% of shape deviations. The relative warps (*i.e*., the principal components or PCs of the Procrustes residuals around the consensus shape) were calculated using the function relWarps from the Morpho package in R (Rohlf 2003, Schlager 2017). The top three relative warps explained 91% of the shape variation in our dataset within alienation space.

#### TPS and statistical tests

The extent to which each specimen-year differs from the consensus shape can be visualized using *thin-plate splines* or TPS. These images represent the deviation of observed specimen landmarks from the consensus shape and interpolate shape changes that occur between the specimen’s landmark coordinates and the analogous coordinates of the consensus shape (Bookstein 1997, Bookstein 2018).

To assess how the different dimensions of alienation (nature, human, and industry) operated in concert, we conducted a within-subject multiple correlation analyses with U.S. state as the subject to determine (1) whether alienation increased over time along the human alienation axis, and (2) whether a relationship could be discerned across the three sources of agriculture alienation in the period 1970-2000.

The effect of annual variation in alienation space on outbreak size was determined by regressing the relative warp scores on state incidence of salmonella and shigellosis, the two foodborne diseases we tested, using MANCOVA. To test for scale effects–at which geographic scales salmonella and shigellosis may correlate with our agriculture alienation shapes–we also tested for significant associations with the partial warps scores and the Procrustes residuals. The latter was conducted using the function procD.lm from the geomorph package with a resampling-based assessment of predictive ability (Adams et al. 2019). The raw data used for these analyses, as well as the accompanying R scripts, are available as a git repository at: https://github.com/kewok/AgriculturalAlienationScripts.

## Results

### U.S. state-specific deviations from the mean alienation configuration over time

By the thin-plate splines shown in Figure 2, we can track relative shifts among the U.S. states across the agriculture alienation space. In particular, we find uneven regime shifts in industrial production. Peak crisis year 1985 approximates the consensus configuration. We see membership in the cutting edge of industrial production in the upper-right change. By 1980, North Dakota has joined the Corn Belt states of Iowa, Illinois, and Indiana in its mix of commodity crops, a shift that Lin and Huang (2019) tracked down to the county level. Its neighbor South Dakota approaches this first tier but by a different trajectory. Other states take the long way toward such production. Colorado on the *y*-axis appears to exit out of labor-intensive production as happens in California, but only Montana trends towards more intensive, fertilizer- and pesticide-led production. Indeed, by 1990, Montana begins to pull away from a second group—Michigan, North Carolina, and Wisconsin. Also on this axis, Missouri escapes Alabama and Michigan falls out of the Upper Midwest group.

**Figure 2.**
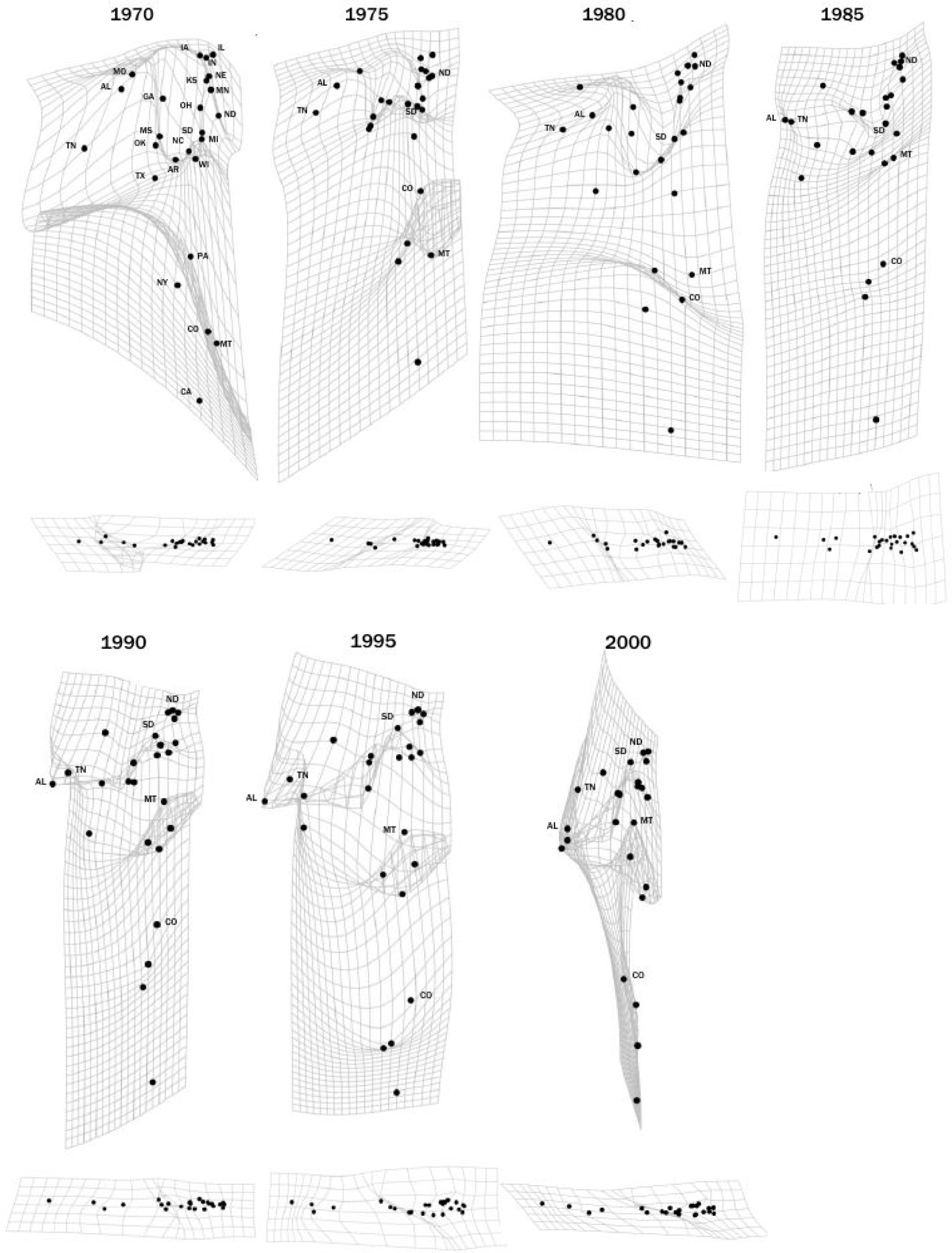
Changes in agricultural alienation space for 25 U.S. states, 1970-2000, depicted as thin-plate splines illustrating how each year’s configuration in alienation space deviates from the consensus shape. For each year, the top spline illustrates deviation along the nature-human alienation axes, and the bottom spline depicts deviation along the human-industry alienation axes.

By 1985 on the nature axis, Tennessee, the last of the study states with considerable cropland pasture, surpasses Alabama converting to commodity annuals, catching up to Texas and Oklahoma. By 2000, we see intimations of what would be, outside specific counties (Baxter and Colvert 2017), the effective end of cropland pasture by 2015, for which the TPS would compress to a near vertical line. Along the industry axis in the bottom TPS, the relative positions across the states appear largely maintained even in and out of the spike in industry alienation of the 1980s crisis, although there is some spread for 1985.

### Key trends in overall shape configuration across time

Table 1 shows the results of our within-subject multiple correlation analyses. With individual U.S. states treated as subjects, in the absence of effects from industrial and natural alienation, human alienation decreases over time. At the same time, when nature alienation increases, human alienation concomitantly increases. Human alienation also increases in concert with industrial alienation over time. Nevertheless, *within any given year*, greater industrial alienation is associated with reduced human alienation. We can summarize the complex of relationships across nature, human, and industry alienation as:

**Table 1.** The linear mixed effect model describing human alienation over time as a function of nature and industry alienation, treating U.S. states as subjects and with year-over-year temporal autocorrelation. Although nature alienation’s effect on human alienation appears non-significant, we retain it here as the linear mixed effect model including nature alienation has a lower Akaike Information Criterion score than either the model excluding nature alienation or the model with a nature alienation ⨯ year interaction effect. We note there is unlikely to be collinearity here, as regressing nature alienation on industry alienation and an interaction term between industry alienation and year (with U.S. state treated as subject and in the presence of temporal autocorrelation) shows no relation between nature and industry alienation or between nature and industry alienation ⨯ time (*p*-value > 0.95).

| Variable | Effect on human alienation | Std. error | df | P-value |
| --- | --- | --- | --- | --- |
| Intercept | 3801.62 | 2249.90 | 146 | 0.093 |
| Nature alienation | 0.61 | 0.34 | 146 | 0.078 |
| Year | -1.93 | 1.13 | 146 | 0.091 |
| Industry alienation | -263.20 | 107.36 | 146 | 0.015 |
| Industry alienation X Year | 0.133 | 0.05 | 146 | 0.015 |

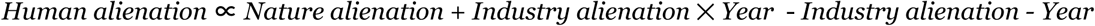

We found no apparent trend in the relationship between nature alienation and industry alienation unmediated by human alienation.

Temporal autocorrelation in human alienation was confirmed by comparing two versions of the linear-mixed effect model in Table 1: one model with temporal autocorrelation and another model without (likelihood-ratio test *p*-value <10^−5^).

Figure 3 shows the relative warps for our ag alienation space for our 25 U.S. states, every five years, 1970-2000. The first two relative warps account for 81% of the total variance across the three alienation axes and appear to recapitulate our qualitative interpretation of Figure 2. RW2 captures the shift in and out of the 1980s crisis. RW1, accounting for 70% of the total variance, shows that switchback leaves the U.S. alienation space in a new configuration.

**Figure 3.**
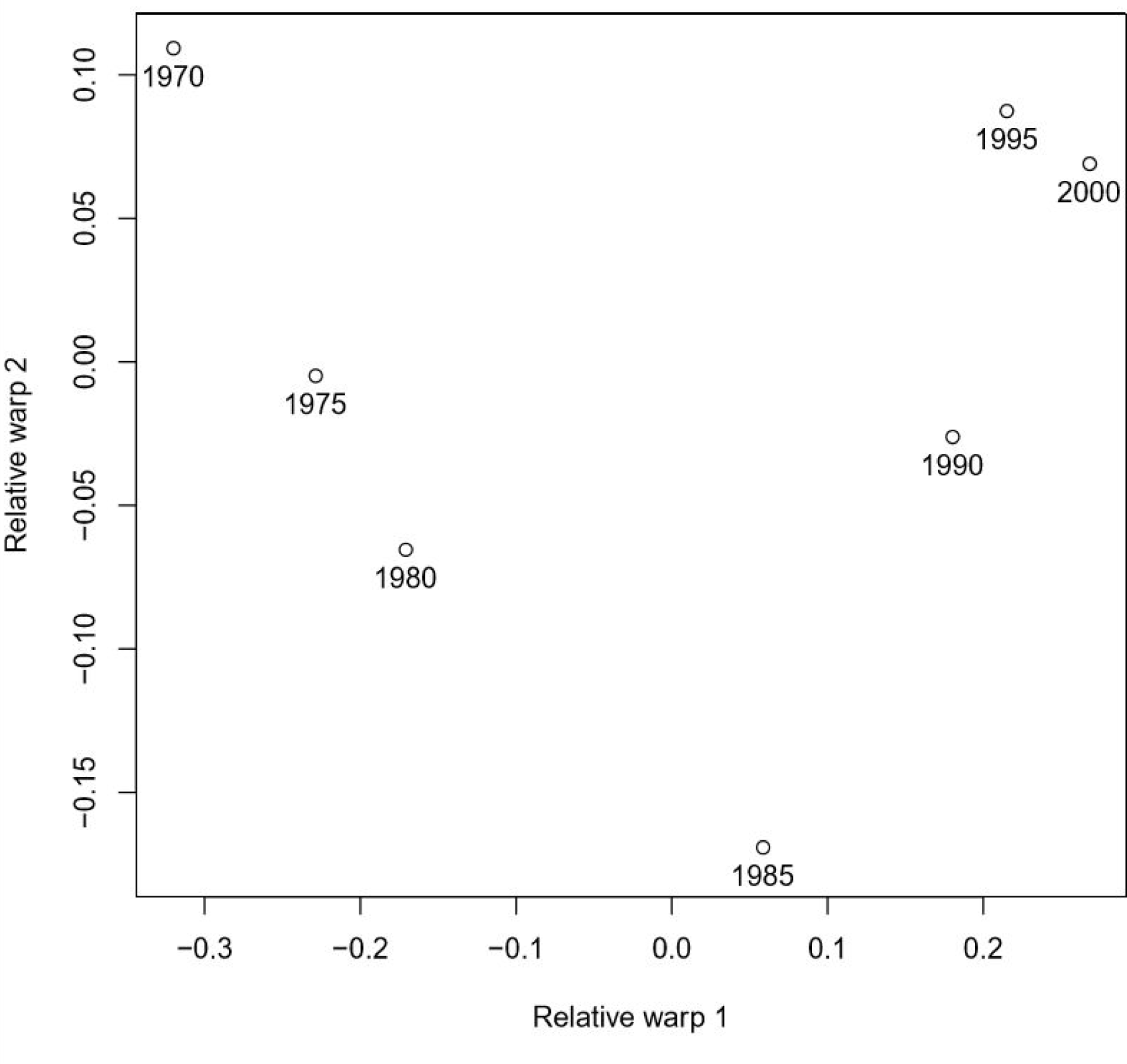
Relative warp scores for each U.S. specimen-year along relative warps 1 and 2, 1970-2000.

### Annual shape configuration as a predictor of outbreak incidence

Table 2 shows annual variation in the national incidences of shigellosis and salmonellosis, the two foodborne illnesses we tested, is generally not associated with alienation shape. Only relative warp 2 marking the farming crisis proved significantly correlated to salmonella outbreaks, although Bonferroni-correcting the resulting *p*-value (0.04) to account for the regressions conducted on the two pathogens suggests relative warp 2’s effect on salmonella incidence to be limited (Bonferroni-corrected *p*-value = 0.09). The scope of the regressions was also limited by sample size, *n* = 7 specimen-years.

**Table 2.** Multiple regressions of salmonella and shigellosis outbreaks on relative warp scores (** uncorrected *p*-value < 0.001; *uncorrected *p*-value < 0.05).

| Variable | Effect on salmonella incidence | Std. Error | Effect on shigellosis incidence | Std. Error |
| --- | --- | --- | --- | --- |
| Intercept | 16.33** | 1.19 | 10.04 | 2.20 |
| Relative warp 1 | 12.35 | 5.42 | 10.18 | 10.00 |
| Relative warp 2 | -43.55* | 13.08 | 0.448 | 24.16 |
| Relative warp 3 | 18.49 | 14.44 | -1.28 | 26.68 |

Regressions for partial warps, encapsulating shape vectors by locale scale, from most local combinations of U.S. states to most general across all 25 states, also showed no significant results following a multiple comparisons correction (Table 3). Likewise, regressing yearly incidence on each year’s Procrustes residuals using permutation tests yielded no significant association between deviation from the consensus shape and outbreak size.

**Table 3.**
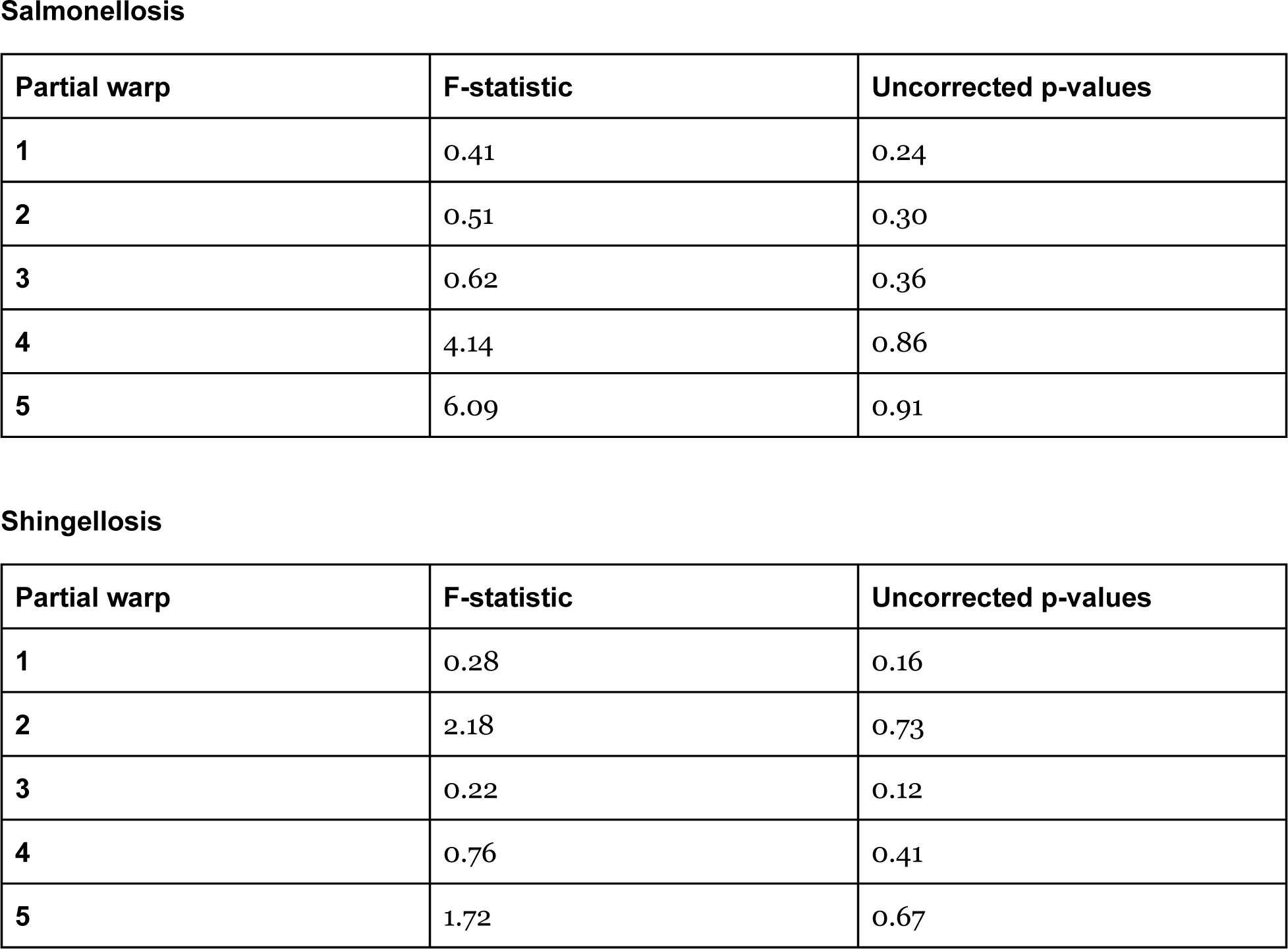
Regression F statistics for the five most impactful U.S. agriculture alienation partial warp variables on salmonellosis and shigellosis incidences reported every five years, 1970-2000. For all regressions, the degrees of freedom associated with each F-statistic are 3,3.

## Discussion

### Capital-led alienation in agriculture

Our analysis of a near-continental agricultural system across 25 U.S. states, 1970-2000, paints a picture of long-term changes in nature, human, and industry alienation in intensive agriculture. Figures 2 and 3 show the system of states undergoing a spike in alienation across all three axes associated with the farm crisis of the early-to-mid 1980s that upon resolution placed the system in a different part of the parameter space: little cropland pasture at the state level, more debt load, and comparatively less money spent on wages. Changes across the study period simultaneously represent an uneven regime shift in agricultural production, with some states holding largely steady in the alienation space and others surging toward a more industrial combination. For instance, by different trajectories North Dakota, South Dakota, and Montana produce localized folds in the thin-plates splines of Figure 2.

Two caveats mark the study. First, the analysis focused on cropland pasture alone. While precipitous declines are apparent across the study states, and indeed all fifty U.S. states, pasture beyond cropland *increased* nationally, largely accounted for by *abandoned* cropland (as opposed to expensive conversions to grassland pasture and range) (Baxter and Calvert 2017). New pasture may represent rewilding by neglect in some counties. In others it may represent little more than a foothold for the real estate bubbles of spreading suburbanization (Tumber 2019). Alongside the difference between permanent and annual pasture, our analysis sidesteps the distinction.

Complex geographies appear at play. U.S. rural areas are neither merely nature bent to human need, nor is their regional interplay the result of central planning. U.S. agriculture has long left behind the strictly teleological modes of increasing cropland acreage *and* the contracted land use brought about by more efficient production (RG Wallace 2018a,b). A highly differentiated landscape is defined unevenly by the dictates of capitalist production operating across multiple commodities, including land itself, even in the face of agribusiness’s widespread substitutionism (Smith 2008, Eaton 2011, Weber and Key 2014). Each sector is actualized in interaction with a confluence of socio-natural factors, among them area-specific legacies in investment, available fixed capital, demographics, soil types, and other historically and regionally contingent factors. Land abandonment *and* increases in overall yield can mark a place entrained by a variety of commercial logics, belying the traditional urban-rural divide. Intensification, underdevelopment, and socio-ecological abandonment can co-exist across a polarized landscape that instantiates multiple momenta in accumulation originating in capital centers near and far (Harvey 1982 [2006], Bergmann and Holmberg 2016, Harvey 2018).

A second caveat is more acutely methodological. Although the relative warps are calculated to maximize orthogonality, Figure 3 suggests a non-monotonic relationship between relative warps 1 and 2. Hence, the full extent of geographic and temporal variability in alienation space is only encapsulated to a first approximation by the principal component analyses. Because we aim to capture the behavior of a large-scale system acting over millions of people, hundreds of thousands of square miles, and multiple ecosocial domains, our results inevitability simplify a complex socioecological topology (Levins and Wilson 1980, Levins 2006).

Despite the linearized simplification, we were able to elucidate complex interactions. Nature alienation is correlated with human alienation. As cropland pasture is converted into commodity crops, expenditures rise in relation to wages. Cropland conversion is tied to increases in farm expenditures on inputs (Hertel and Baldos 2016). Pasture, while generally producing less in direct revenue than cropland (albeit a trend in some regions now in reverse), also demands significantly less in fertilizer, seeds, and herbicides per acre (Lacefield and Ball 2015). Increases in expenditures on inputs drain the on-farm cash value derived from crop production (and by extension livestock) (Begemann 2017, USDA-Economic Research Service 2019b).

That revenue capture instantiates broader political economies. U.S. rural landscapes (and adjunct ethoses) long associated with reproducing community life generation-to-generation well beyond the farm gate have been largely transformed into mosaics of individualized economic units organized around reproducing suburbanized agribusiness first (Berry 2015, Meyer and Graybill 2016, Ramsey 2016). The local ecosystems and human communities upon which this agricultural extractivism depends are treated as sacrifice zones for spatially externalized sources of capital accumulation (Weber 2018, Edelman 2019).

Table 1 further shows an inverse relationship between human alienation and industrial alienation, even as, within any given year, the two variables are directly related. Thus, across longer time horizons, as debt/equity ratios decrease, there is a smaller gap between farmer wages and fertilizer (and other) expenditures. Superficially, this may indicate that industrial alienation can, in fact, reduce human alienation. But there may be a more troubling dynamic in the mix. The outcome may reflect the extent to which farmers take on debt under the corporate food regime to pay themselves on-farm wages, as well as, outside this analysis, their farmhands (Bergman et al. 2017, Graddy-Lovelace and Diamond 2017).

Farmers may be leveraging their equity and assets, including their land and machinery, to pay themselves at the expense of both their autonomy and capacity to socially reproduce themselves beyond the short term (Rotz and Fraser 2015). Long-term trends in a generational process of immiseration may be obscured if wages are paid for or floated by passing cash injections smoothing acute market and climate-led weather shocks (Bergman et al. 2017). Table 1 supports the dual-dynamic of a financial *performance* that stabilizes or even surges year-to-year as farmers’ financial *positions* deteriorate (USDA-Economic Research Service 2019a).

The bifurcation manifests in social geography. With so much of their revenue structurally dedicated to overcapitalizing on inputs and, at the other end of production, settling for end buyers’ monopolistic pricing, even the more successful U.S. farmers, many of whom survived by consolidating their neighbors’ farms into economies of scale, are still relying on loans, federal crop insurance, fixed direct payments, and off-farm income (Burns and Prager 2016, Bekkerman et al. 2019). Much as any semi-proletarianized farmer or rural precariat of the global South, or itinerant migrant labor they hire stateside, U.S. farmers, albeit inside a different risk matrix, are cycle-migrating from farm to urban employment or even in the same spot working remotely (Arrighi 2009, Bryceson 2010, Baird 2011, Hornby et al. 2018). This precarity undercuts the stable sociality needed to organize resistance, or an out-and-out alternative, to the agribusiness food regime (Arrighi 2009, Davis 2018). The declining financial position also selects for selling off farmland, by far farmers’ main asset, to suburban sprawl or high-end investors looking to diversify their portfolios (Fairbairn 2014, Tumber 2019).

### Diseases can mark and make alienation

Pathogens can amplify the very capital-led alienation that selected for them in agriculture. Industrial conditions select for disease virulence, transmissibility, and genomic reassortment (Nelson et al. 2015, Rozins and Day 2017, Dhingra et al. 2018). With poultry and livestock breeding conducted offshore for economically lucrative morphometric characteristics and reproduction impossible on-site, no immune resistance found in an outbreak’s survivors can be passed on to the next generation (RG Wallace 2016a). To insure pathogens do not spread within and across pig herds, specific-pathogen-free offspring are produced by terminal hysterectomy (RG Wallace et al. in preparation). Sow uteri are removed and placed in humidicribs or doused in antiseptic before the piglets are removed from their uterus casing. Piglets are then isolated and some medically induced into early weaning. By such path dependencies, biosecurity disconnects livestock and poultry from their natural environments and sociality. Farmers must pay extra for these specially produced cohorts, and smallholders are punished for failing to follow biocontrol protocols for diseases of industrial sourcing (Bryant and Garnham 2014, RG Wallace 2018c).

The resulting alienation extends to scientific study itself. Our attempts at quantifying the epidemiological effects of shifts in alienation space here are constrained by the few recorded and publicly reported infections that have steadily circulated across U.S. poultry and livestock across the study period. Among hog, for instance, African swine fever has never been reported in the U.S., although an ongoing global outbreak may soon change matters (Brown and Bevins 2018). Classic swine fever was eradicated in 1978. Swine brucellosis and pseudorabies are nowhere detected in commercial herds. Some of the more serious reportable infections are only recent U.S. introductions, among them porcine reproductive and respiratory syndrome virus in 1987, its highly pathogenic variant in the mid-2000s, and porcine epidemic diarrhea virus in 2013 (Mole 2013, Lunney et al. 2016).

Influenza offers one exception. Swine influenzas have been recorded at least as far back as infection with the 1918 human strain, changing little until the Livestock Revolution restructured the hog sector (RG Wallace 2009). A phylodynamics based on influenza genetic sequences may be able to reconstruct the effective population size of H1 strains in hog back to the start of our sample period, even under the pressure of fewer samples earlier on. Nelson et al.’s (2011) reconstruction of the swine origins of H1N1 (2009) extends only as far back as 2004. Lin et al. (2011) estimate effective flu population size based on genetic differences back before 1970 (although only from 1998 do the estimates exhibit seasonal dynamics).

There are complications attached to such a research prospectus. Anderson et al. (2013) report 1000+ genetic sequences collected 2009-2012 across the U.S. for each of three swine influenza genomic segments for strains A/H1N1, A/H3N2, and A/H1N2. The Influenza Research Database lists over 3000 partial and full influenza hemagglutinin sequences for U.S. swine, with 123 of those sequences before 2000, less densely populated back in time. But much of the reporting system depends on permission from hog producers and the U.S. Centers for Disease Control and the Department of Agriculture units catering to their good will (Nelson et al. 2011, RG Wallace 2016b). Other independent researchers as yet unaffiliated with the reporting program may be blocked access to many of the samples. That is, the data themselves may be a manifestation of agricultural alienation (Goe and Kenney 1988, Shill 1992, Evans 1992, Van Dijk et al. 2010).

### Political epistemology

Complications extend beyond data availability. The pertinent methodological issue at hand is often less a matter of the statistical model to be used, however important such choices remain. The key epistemological quandary can be found in the underlying normative presumptions surrounding, in this case, agricultural production (R Wallace et al. 2018). Investigators must specify *a priori* which questions are to be studied, decisions oft-tied to political economies found far afield (Levins 1998). Such ghosts are also found in the modeling machine. Even under the kind of machine learning that parses through combinatorials of candidate variables automatically, the resulting work can find sociopolitical norms folded in as far down as model formalisms and their operationalization long before any end-use application (Levins 1998, 2006, Winther 2006, RG Wallace 2011, Schizas 2012, Hajian et al. 2016, Nikisianis and Stamou 2016). Including only one of the three types of alienation is insufficient as a matter of both method and practice for most of the kinds of study systems we address here. The resulting modes of failure in analysis are likely to extend to interventions, exasperating the very problems the work aims to alleviate.

For instance, many of the growing number of agro-circular economic models aim at reconnecting nature to the neoliberal economy. The economies typically proposed in the stead of disembedded production presume linear biophysical cascades can be recoupled by recycling product and waste alike (*e.g*., Yuan et al. 2006, Xuan et al. 2011, Jun and Xiang 2011, Heshmati 2015, Quina et al. 2017, Govidan and Hasanagic 2018). As is the case in much of green capitalism, production efficiencies are proposed to minimize energy use and raw materials (Foster and Burkett 2016). Research around circular economies appears largely qualitative and aspirational at this point. Consumer surveys are measuring consumer acceptability of recycled byproducts (Borello et al. 2017).

As statistical studies of the metabolic rift are in their own infancy, we need not harry these efforts on account of their early stage. But models in circular economy are often also presently framed in entrepreneurial terms, leaving out the contraindicative Jevons paradox against production efficiencies (Jevons 1865, Foster et al. 2010, RG Wallace and Kock 2012). Under the paradox, technological innovations cheapen production, helping open new markets for raw resources. As a result, more resources are used and more of the environment damaged. The issue extends beyond such perverse outcomes. In promoting large-scale ecological change, many environmental modelers presume, even celebrate, continuing privatization of expropriated surplus value (*e.g*., Nassauer et al. 2002, Jordan et al. 2016). The ethos is repeatedly operationalized as if the natural order of things. Entering nature into the market, “selling nature to save it,” is treated as an unspoken social good (Robertson 2006, Lansing 2013, Dempsey and Suarez 2016). Irreplaceable “ecosystem services” are proposed to be monetizable to the benefit of the very capital destroying them. Against such claims, the standard operation procedures of agribusiness power are locked in together. Feed-the-world narratives, compartmentalized thinking, export economics, measures of success in terms of productivity and yield alone, land grabbing, externalizing damage to land and labor alike, and structural and State violences to enforce these costs upon alternate food regimes are irreducibly bounded (Patel 2013, IPES-Food 2016, R Wallace et al. submitted).

The export economies these putative remedies support spirit social surplus offshore, driving the very ecosystemic degradation the new models ostensibly aim to remediate (Perfecto and Vandermeer 2010, Bergmann and Holmberg 2016). The broader political economy compounds such unequal ecological exchange (Freebairn 1995, Jorgenson 2006, Chappell et al. 2013). Failures to keep up with the race-to-the-bottom of international competition are punished, even in the U.S system studied here as elsewhere across the global North. Less extractive regimens, or, in a producer-led economy, target markets that refuse to accept goods produced out of the basest extractivism are simply replaced (Oliveira and Schneider 2016). So multinational producers ride lucrative spatial fixes, logging forest to forest and region to region (Harvey 1982 [2006], Song et al. 2015). In desperation, the poor slash-and-burn beyond traditional schedules on the marginal land to which they are confined (Ioris 2015, Sauer 2018). Additional offsets are imposed by a global financialization that treats cash crops and food animals as living fungibilities with commitments beyond mere commoditization months before their germination or birth (RG Wallace et al. in preparation). Species are treated as asset classes subject to price volatilities favored as ontologically realer than the ecological or epidemiological dynamics into which the non-human stock will eventually be grown.

In this context, the metabolic rifts of global impact with which we began the Introduction can be solved only across all three alienation axes simultaneously. Agroecological interventions must extend beyond the biophysical (Altieri et al. 2017, Perfecto et al. 2019, Giraldo 2019). Farmer autonomy and a strong public sector can together curb environmental ratchets and runaway infections, wherein diseases routinely marginalized to local outbreaks suddenly surge to regional and even protopandemic threats (R Wallace et al. 2018). Even such efforts are insufficient without a means by which to defend these experiments from the compulsions neoliberal economics impose upon individuals and communities alike or the threat of capital-led State repression (R Wallace et al. submitted).

In our attempt at correlating a complex, agro-socioeological landscape to sparsely populated incidence data, we did not find a linear correlation between systematic alienation and the two foodborne diseases we tested. Nevertheless, our analyses may offer one kind of modeling framework for including such broader structural factors rather than letting these variables be zeroed out by expedient economism (RG Wallace et al. 2015, R Wallace et al. 2018). Researchers should consider folding in the accumulating evidence that successful epidemiological interventions are predicated upon a robust social commons (Mutaner et al. 2000, Pellizzoni 2018). Statistical analyses need not be as alienated as the subjects they address. Another science is possible.

## Data Availability

The raw data used for these analyses, as well as the accompanying R scripts, are available as a git
repository at: https://github.com/kewok/AgriculturalAlienationScripts.

## Literature Cited

Adams DC, ML Collyer, and A Kaliontzopoulou (2019) Geomorph: Software for geometric morphometric analyses. R package version 3.1.0. https://cran.r-project.org/package=geomorph.

Alexandratos N and J Bruinsma (2012) World Agriculture Towards 2030/2050: The 2012 Revision. ESA Working Paper 12-03. Agricultural Development Economics Division, Food and Agriculture Organization.http://www.fao.org/fileadmin/templates/esa/Global_persepctives/world_ag_2030_50_2012_rev.pdf.

Allen CD, DD Breshears, and NG McDowell (2015) On underestimation of global vulnerability to tree mortality and forest die-off from hotter drought in the Anthropocene. Ecosphere, 6(8). https://doi.org/10.1890/ES15-00203.1

Altieri MA, CI Nicholls, and R Montalba (2017) Technological approaches to sustainable agriculture at a crossroads: An agroecological perspective. Sustainability, 9(3):349.

Anderson TK, MI Nelson, P Kitikoon, SL Swenson, JA Korslund, and AL Vincent (2013) Population dynamics of cocirculating swine influenza A viruses in the United States from 2009 to 2012. Influenza Other Respir Viruses, 7(S4):42–51.

Anderson TM, et al. (2018) Herbivory and eutrophication mediate grassland plant nutrient responses across a global climatic gradient. Ecology, 99(4):822–831.

Arrighi G (2009) The winding paths of capital: Interview by David Harvey. New Left Review, 56:61–94.

Baer H and M Singer (2016) Global Warming and the Political Ecology of Health. Routledge, London.

Baird IG (2011) Turning land into capital, turning people into labour: Primitive accumulation and the arrival of large–scale economic land concessions in the Lao People’s Democratic Republic. New Proposals: Journal of Marxism and Interdisciplinary Inquiry, 5:10–26.

Barnosky AD, et al. (2012) Approaching a state shift in Earth’s biosphere. Nature, 486:52–58.

Baxter RE and KE Colvert (2017) Estimating available abandoned cropland in the United States: Possibilities for energy crop production. Annals of the American Association of Geographers, 107(5):1162–1178.

Begemann S (2017) Seed price triples over last 20 years. AgWeb, 20 July. Available online at https://www.agweb.com/article/seed-price-triples-over-last-20-years-NAA-sonja-begemann.

Bekkerman A, EJ Belasco, and VH Smith (2019) Does farm size matter? Distribution of crop insurance subsidies and government program payments across U.S. farms. Applied Economic Perspectives and Policy, 41(3):498–518.

Bergman NK, R Iyer, and RT Thakor (2017) The Effect of Cash Injections: Evidence from the 1980s Farm Debt Crisis. National Bureau of Economic Research. NBER Working Paper No. 23546. Available online at https://www.nber.org/papers/w23546.pdf.

Bergmann LR (2013) Bound by chains of carbon: Ecological-economic geographies of globalization. Annals of the Association of American Geographers, 103:13481370.

Bergmann LR (2017) Towards economic geographies beyond the Nature-Society divide. Geoforum, 85:324–335.

Bergmann LR and M Holmberg (2016) Land in motion. Annals of the American Association of Geographers, 106(4):932–956.

Bookstein FL (1991) Morphometric Tools for Landmark Data. Cambridge University Press, Cambridge.

Bookstein FL (1997) Landmark methods for forms without landmarks: morphometrics of group differences in outline shape. Med. Image Anal. 1:225–243.

Bookstein FL (2018) A Course in Morphometrics for Biologists: Geometry and Statistics for Studies of Organismal Form. Cambridge University Press, UK.

Borrello M, F Caracciolo, A Lombardi, S Pascucci, and L Cembalo (2017) Consumers’ perspective on circular economy strategy for reducing food waste. Sustainability, 9(141). doi:10.3390/su9010141

Brown VR and SN Bevins (2018) A review of African swine fever and the potential for introduction into the United States and the possibility of subsequent establishment in feral swine and native ticks. Frontiers in Veterinary Science, 5: 11.

Bryant L and B Garnham (2017) Glocal terrains of farmer distress and suicide. In M Livholts and L Bryant (eds), Social Work in a Glocalised World. Routledge, New York, pp 25–41.

Bryceson DF (2010) Sub-Saharan Africa’s vanishing peasantries and the specter of a global food crisis. In F Magdoff and B Tokar (eds), Agriculture and Food in Crisis: Conflict, Resistance and Renewal. Monthly Review Press, New York.

Büntgen U, et al. (2011) 2500 years of European climate variability and human susceptibility. Science, 331(6017) 578–582.

Burns CB and DL Prager (2016) Do direct payments and crop insurance influence commercial farm survival and decisions to expand? Selected paper presented for presentation at the 2016 Agricultural & Applied Economics Association, Boston, MA, July 31-August 2. https://ageconsearch.umn.edu/record/235693/files/Burns_Prager_AAEA_2016_Crop_Insurance_Farm_Survival_Expansion.pdf.

Bryant L and B Garnham (2014) Economies, ethics and emotions: Farmer distress within the moral economy of agribusiness. Journal of Rural Studies, 34:304–312.

Carpenter SR, EH Stanley, and MJV Zanden (2011) State of the world’s freshwater ecosystems: Physical, chemical, and biological changes. Annual Review of Environment and Resources, 36:75–99.

Chakrabarty D (2009) The climate of history: Four theses. Critical Inquiry, 35(2):197–222.

Chappell, MJ, H Wittman, CM Bacon, BG Ferguson, LG Barrios, et al. (2013) Food sovereignty: an alternative paradigm for poverty reduction and biodiversity conservation in Latin America. F1000Res, 2:235.

Choat B, et al. (2012) Global convergence in the vulnerability of forests to drought. Nature, 491:752–755.

Clausen R, B Clark, and S.B. Longo (2015) Metabolic rifts and restoration: Agricultural crises and the potential of Cuba’s organic, socialist approach to food production. World Review of Political Economy, 6(1):4–32.

Clement MT (2009) A basic accounting of variation in municipal solid-waste generation at the county level in Texas, 2006: Groundwork for applying metabolic-rift theory to waste generation. Rural Sociology, 74(3):412–429.

Da Costa, P. M., Loureiro, L., & Matos, A. J. (2013). Transfer of multidrug-resistant bacteria between intermingled ecological niches: the interface between humans, animals and the environment. International Journal of Environmental Research and Public Health, 10: 278–294.

Dakos V, B Matthews, AP Hendry, J Levine, N Loeuille, J Norberg, P Nosil, M Scheffer, and L De Meester (2019) Ecosystem tipping points in an evolving world. Nature Ecology & Evolution, 3:355–362.

Daugherty AB (1991) Major Uses of Land in the United States: 1987. Agricultural Economics Report No. 643. Economic Research Service, U.S. Department of Agriculture.

Daugherty AB (1995) Major Uses of Land in the United States, 1992. Agricultural Economics Report No. 723, Economic Research Service, U.S. Department of Agriculture.

Dawson A (2016) Extinction: A Radical History. OR Books, New York.

Dempsey J and DC Suarez (2016) Arrested development? The promises and paradoxes of “selling nature to save it”. Annals of the American Association of Geographers, 106:653–671.

Dhingra MS, J Artois, S Dellicour, P Lemey, G Dauphin, et al. (2018) Geographical and historical patterns in the emergences of novel Highly Pathogenic Avian Influenza (HPAI) H5 and H7 viruses in poultry. Front. Vet. Sci., 05 https://doi.org/10.3389/fvets.2018.00084

Diamond ML, et al. (2015) Exploring the planetary boundary for chemical pollution. Environment International, 78:8–15.

Drijfhout S, S Bathiany, C Beaulieu, V Brovkin, M Claussen, C Huntingford, M Scheffer, G Sgubin, and D Swingedouw (2015) Catalogue of abrupt shifts in Intergovernmental Panel on Climate Change climate models. PNAS, 112(43):E5777–E5786.

Dryden, I. L. (2018). shapes: Statistical Shape Analysis. R package version 1.2.4. https://CRAN.R-project.org/package=shapes

Eaton E (2011) On the farm and in the field: The production of nature meets the Agrarian Question, New Political Economy, 16(2):247–251.

Edelman M (2019) Hollowed out Heartland, USA: How capital sacrificed communities and paved the way for authoritarian populism. Journal of Rural Studies. Available online at https://www.sciencedirect.com/science/article/abs/pii/S0743016719305157.

Ellis EC and N Ramankutty (2008) Putting people in the map: anthropogenic biomes of the world. Frontiers in Ecology and the Environment, 6(8):439–447.

Engering A, L Hogerwerf, and J Slingenbergh (2013) Pathogen-host-environment interplay and disease emergence. Emerging Microbes and Infections, 2:e5.

Essington, TE, et al. (2015) Fishing amplifies forage fish population collapses. Proceedings of the National Academy of Sciences of the United States of America, 112(21):6648–6652.

Evans JF (1992) Issues in equitable access to agricultural information. Agriculture and Human Values. 9(2):80–85.

Fairbairn M (2014) ‘Like gold with yield’: Evolving intersections between farmland and finance. Journal of Peasant Studies, 41(5):777–795.

FAO (2013) World Livestock 2013: Changing Disease Landscapes. Food and Agriculture Organization, United Nations, Rome.

Filippelli GM and MP Taylor (2018) Addressing pollution-related global environmental health burdens. GeoHealth, 2(1):2–5.

Foley J, R Defries, GP Asner, C Barford, G Bonan, SR Carpenter, et al. (2005) Global consequences of land use. Science, 309:570–574.

Foster JB (2016) Marx as a food theorist. Monthly Review, 68(7).

Foster JB and P Burkett (2016) Marx and the Earth: An Anti-Critique. Brill, Leiden.

Foster JB, R York, and B Clark (2010) The Ecological Rift: Capitalism’s War on the Earth. Monthly Review Press, New York

Freebairn DK (1995) Did the Green Revolution concentrate incomes? A quantitative study of research reports. World Dev., 23(2):265–279.

Frey HT (1973) Major Uses of Land in the United States--Summary for 1969. Agriculture Economics Report No. 247. Economic Research Service, U.S. Department of Agriculture.

Frey HT (1979) Major Uses of Land in the United States: 1974. Agriculture Economics Report No. 440. Economics, Statistics, and Cooperatives Service, U.S. Department of Agriculture.

Frey HT (1982) Major Uses of Land in the United States: 1978. Agriculture Economics Report No. 487. Economic Research Service, U.S. Department of Agriculture.

Frey HT and R Hexem (1985) Major Uses of Land in the United States: 1982. Agriculture Economics Report No. 535. Economic Research Service, U.S. Department of Agriculture.

Frey HT, OE Krause, and C Dickason (1968) Major Uses of Land and Water in the United States, Summary for 1964. Agriculture Economics Report No. 149, Economic Research Service, U.S. Department of Agriculture.

Gauthier S, P Bernier, T Kuuluvainen, and AZ Shvidenko (2015) Boreal forest health and global change. Science, 349(6250):819–822.

Giraldo OF (2019) Political Ecology of Agriculture: Agroecology and Post-Development. Springer, Cham.

Goe WR and M Kenney (1988) The political economy of the privatization of agricultural information: The case of the United States. Agricultural Administration and Extension, 28(2):81–99.

Govindan K and M Hasanagic (2018) A systematic review on drivers, barriers, and practices towards circular economy: a supply chain perspective, International Journal of Production Research, 56(1-2): 278–311.

GRAIN and IATP (2018) Emissions Impossible: How Big Meat and Dairy Are Heating up the Planet. https://www.iatp.org/emissions-impossible.

Groseclose SL, WS Brathwaite, PA Hall, CM Knowles, D. Adams, et al. (2003) Summary of notifiable diseases -- United States, 2001. MMWR Weekly, 50(53):1–108.

Gunderson R (2011) The metabolic rifts of livestock agribusiness. Organization & Environment, 24(4):404–422.

Hajian, S., Bonchi, F., & Castillo, C. (2016). Algorithmic bias: From discrimination discovery to fairness-aware data mining. In Proceedings of the 22nd ACM SIGKDD international conference on knowledge discovery and data mining, ACM pp. 2125-2126.

Hamilton HA, D Ivanova, K Stadler, S Merciai, J Schmidt, R van Zelm, D Moran, and R Wood (2018) Trade and the role of non-food commodities for global eutrophication. Nature Sustainability, 1:314–321.

Harvey D (1982 [2006]) The Limits to Capital. Verso, New York.

Harvey D (1993 [2016]) The nature of environment: The dialectics of social and environmental change. In The Ways of the World. Oxford University Press, Oxford, pp 159–213.

Harvey D (2018) Marx’s refusal of the labour theory of value: Reading Marx’s Capital with David Harvey. 14 March.http://davidharvey.org/2018/03/marxsrefusal-of-the-labour-theory-of-value-by-david-harvey/

Heede R (2014) Tracing anthropogenic carbon dioxide and methane emissions to fossil fuel and cement producers, 1854–2010. Climatic Change, 122(1–2):229–241.

Hertel TW and ULC Baldos (2016) Productivity growth and yields in the global crops sector. In Global Change and the Challenges of Sustainably Feeding a Growing Planet. Springer, Cham,, pp 27–39.

Heshmati, A (2015) A Review of the Circular Economy and its Implementation. IZA Discussion Papers, No. 9611, Institute for the Study of Labor (IZA), Bonn. https://www.econstor.eu/bitstream/10419/130297/1/dp9611.pdf.

Hornby D, A Nel, S Chademana, and N Khanyile (2018) A slipping hold? Farm dweller precarity in South Africa’s changing agrarian economy and climate. Land, 7(2):40.

Hughes TP, S Carpenter, J Rockström, M Scheffer, and B Walker (2013) Multiscale regime shifts and planetary boundaries. Trends in Ecology and Evolution, 28(7):398–395.

Intergovernmental Panel on Climate Change (2014) AR5 Climate Change 2014: Impacts, Adaptation, and Vulnerability. https://www.ipcc.ch/report/ar5/wg2/

Intergovernmental Panel on Climate Change (2018) Special Report on Global Warming of 1.5 °C (SR15).http://www.ipcc.ch/report/sr15/.

Intergovernmental Panel on Climate Change (2019). Climate Change and Land. https://www.ipcc.ch/site/assets/uploads/2019/08/Fullreport-1.pdf

Ioris AAR (2015) The production of poverty and the poverty of production in the Amazon: Reflections from those at the sharp end of development. Capitalism Nature Socialism, 26(4):176–192.

IPES-Food (2016) From Uniformity to Diversity: A Paradigm Shift from Industrial Agriculture to Diversified Agroecological Systems. Louvain-la-Neuve, Belgium. http://www.ipes-food.org/_img/upload/files/UniformityToDiversity_FULL.pdf.

Jevons WS (1865) The Coal Question: An Inquiry Concerning the Progress of the Nation, and the Probable Exhaustion of Our Coal Mines. Macmillan, London.

Jones BA, D Grace, R Kock, S Alonso, J Rushton, MY Said, et al. (2013) Zoonosis emergence linked to agricultural intensiffcation and environmental change. PNAS, 110:8399–8404.

Jordan N., CS Slotterback, D Mulla, and L Kne (2017) Agriculture and the river: The university’s role in societal learning, innovation, and action. Open Rivers: Rethinking The Mississippi, 6:61–71. http://editions.lib.umn.edu/openrivers/article/agriculture-and-the-river-the-universitys-role-in-societal-learning-innovation-and-action/

Jorgenson AK (2006) Unequal ecological exchange and environmental degradation: a theoretical proposition and cross-national study of deforestation, 1990-2000. Rural Sociology, 71:685–712.

Jun H and H Xiang (2011) Development of circular economy is a fundamental way to achieve agriculture sustainable development in China. Energy Procedia, 5:1530–1534.

Kolbert E (2014) The Sixth Extinction: An Unnatural History. Bloomsbury Publishing.

Lacefield G and D Ball (2015) Stakeholders integration for sustainable use of temperate forage/livestock agriculture. Proceedings of 23rd International Grassland Congress 2015-Keynote Lectures. Available online at https://uknowledge.uky.edu/cgi/viewcontent.cgi?article=1032&context=igc.

Levins R (1998) The internal and external in explanatory theories, Science as Culture, 7(4):557–582.

Levins R (2006) Strategies of abstraction, Biol Philos, 21:741–755.

Levins R and C Lopez (1999) Toward an ecosocial view of health. International Journal of Health Services, 29(2):261–293.

Levins R and M Wilson (1980) Ecological theory and pest management. Annual Review of Entomology, 25(1):287–308.

Lewontin R (1998) The maturing of capitalist agriculture: Farmer as proletarian. Monthly Review, 50(3):72.

Lin JH, SC Chiu, JC Cheng, HW Chang, KL Hsiao, et al. (2011) Phylodynamics and molecular evolution of influenza A virus nucleoprotein genes in Taiwan between 1979 and 2009. PLoS One, 6(8):e23454.

Lin M and Q Huang (2019) Exploring the relationship between agricultural intensification and changes in cropland areas in the US. Agriculture, Ecosystems & Environment, 274:33–40.

Lowder SK, J Skoet, and T Raney (2016) The number, size, and distribution of farms, smallholder farms, and family farms worldwide. World Dev., 87:16–29

Lubowski R, S Buchotlz, R Claassen, MJ Roberts, JC Cooper, A Gueorguieva, and R Johansson (2006) Environmental Effects of Agricultural Land-Use Change: The Role of Economics and Policy. Economic Research Report Number 25. Economic Research Service, United States Department of Agriculture.

Lunney JK, Y Fang, A Ladinig, N Chen, Y Li, B Rowland, GJ Renukaradhya (2016) Porcine reproductive and respiratory syndrome virus (PRRSV): pathogenesis and interaction with the immune system. Annual Review of Animal Biosciences, 4:129–154.

Mac Nally R, et al. (2009) Collapse of avifauna: Climate change appears to exacerbate habitat loss and degradation. Diversity and Distributions, 15(4):720–730.

Malm A (2016) Fossil Capital: The Rise of Steam Power and the Roots of Global Warming. Verso, New York.

Malm A and A Hornborg (2014) The geology of mankind? A critique of the Anthropocene narrative. The Anthropocene Review, 1(1):62–69.

Mancus P (2007) Nitrogen fertilizer dependency and its contradictions: A theoretical exploration of social-ecological metabolism. Rural Sociology, 72(2):269–288.

Mateo-Sagasta J, SM Zadeh, and H Turral (2017) Water Pollution from Agriculture: A Global Review.

The Food and Agriculture Organization and the International Water Management Institute. FAO, Rome.

McClintock N (2010) Why Farm the City? Theorizing Urban Agriculture Through a Lens of Metabolic Rift. Urban Studies and Planning Faculty Publications and Presentations. 91. https://pdxscholar.library.pdx.edu/usp_fac/91.

McGee JA and C Alvarez (2016) Sustaining without changing: The metabolic rift of certified organic farming. Sustainability, 8(2):115.

McIntosh, RJ, JA Tainter, and SK McIntosh (eds) (2000) The Way the Wind Blows: Climate, History, and Human Action. Columbia University Press, New York.

Mészáros I (1970) Marx’s Theory of Alienation. Merlin Press, UK.

Meyer WB and JK Graybill (2016) The suburban bias of American society? Urban Geography, 37(6):863–882.

Mole B (2013) Deadly pig virus slips through US borders. Nature, 499(7459):388.

Moore J (2008) Ecological crises and the agrarian question in world-historical perspective. Monthly Review, 60(6).

Moore J (2015) Capitalism in the Web of Life: Ecology and the Accumulation of Capital. Verso, New York.

Moore J (2017) The Capitalocene, Part I: on the nature and origins of our ecological crisis. Journal of Peasant Studies, 44(3): 594–630.

Muntaner C, J Lynch, and GD Smith (2000) Social capital and the third way in public health. Critical Public Health, 10(2):107–124.

Myers, SS, et al. (2014) Increasing CO2 threatens human nutrition. Nature, 510:139–142.

Nassauer JI, RC Corry, and RM Cruse (2002) The landscape in 2025: alternative future landscape scenarios: a means to consider agricultural policy. Journal of Soil and Water Conservation, 57(2):44A–53A.

Nelson MI, P Lemey, Y Tan, A Vincent, TT-Y Lam, S Detmer, et al. (2011) Spatial dynamics of human-origin H1 influenza A virus in North American swine. PLoS Pathog, 7(6): e1002077.

Nelson MI, C Viboud, AL Vincent, MR Culhane, SE Detmer, et al. (2015) Global migration of influenza A viruses in swine. Nature Communications, 6:6696.

Nickerson C, R Ebel, A Borchers, and F Carriazo (2011). Major Uses of Land in the United States, 2007.

Economic Information Bulletin Number 89. U.S. Department of Agriculture, Washington DC.

Nikisianis N and GP Stamou (2016) Harmony as ideology: Questioning the diversity-stability hypothesis, Acta Biotheoretica, 64(1):33–64.

O’Hara PA (2009) Political economy of climate change, ecological destruction and uneven development. Ecological Economics, 69(2):223–234.

O’Sullivan, D., L, Bergmann, and J.E. Thatcher (2017) Spatiality, maps, and mathematics in critical human geography: Toward a repetition with difference. The Professional Geographer, 70(1):129–139.

Oliveira GLT and M Schneider (2016) The politics of flexing soybeans: China, Brazil and global agroindustrial restructuring. Journal of Peasant Studies, 43(1):167–194.

Patel R (2013) The Long Green Revolution. The Journal of Peasant Studies, 40(1):1–63.

Pellizzoni L (2018) Joining people with things. The commons and environmental sociology. In M Boström and DJ Davidson (eds), Environment and Society: Concepts and Challenges. Springer, Cham, pp 281–304.

Peres CA, T Emilio, J Schietti, SJ. Desmoulière, and T Levi (2016) Dispersal limitation induces long-term biomass collapse in overhunted Amazonian forests. Proceedings of the National Academy of Sciences of the United States of America, 113(4):892–897.

Perfecto I and J Vandermeer (2010) The agroecological matrix as alternative to the land-sparing/agriculture intensification model. Proceedings of the National Academy of Sciences, 107(13):5786–5791.

Perfecto I, J Vandermeer, and A Wright (2019) Nature’s Matrix: Linking Agriculture, Biodiversity Conservation and Food Sovereignty. 2nd Edition. Easthscan, London.

Perry CT and KM Morgan (2017) Bleaching drives collapse in reef carbonate budgets and reef growth potential on southern Maldives reefs. Scientific Reports, 7:40581

Pereira HM, LM Navarro, and IS Martins (2012) Global biodiversity change: the bad, the good, and the unknown. Annu. Rev. Environ. Resourc., 37:25–50.

Peters DJ, SM Monnat, A. Hochstetler, and MT Berg (2019) The opioid hydra: Understanding overdose mortality epidemics and syndemics across the rural-urban continuum. Rural Sociology. https://doi.org/10.1111/ruso.12307.

Piketty T (2014) Capital in the Twenty-First Century. Belknap Press, Cambridge, MA.

Potts SG, et al. (2010) Global pollinator declines: trends, impacts and drivers. Trends in Ecology & Evolution, 25(6):345–353.

Quina MJ,MAR Soares, and R Quinta-Ferreira (2017) Applications of industrial eggshell as a valuable anthropogenic resource. Resources, Conservation and Recycling, 123:176–186.

Ramankutty N, Z Mehrabi, K Waha, L Jarvis, C Kremen, M Herrero, and LH Rieseberg (2018) Trends in global agricultural land use: Implications for environmental health and food security. Annu Rev Plant Biol., 69:789–815.

Ramsey EF (2016) From Cow Pasture to Cul-de-sac: The Intersection of Rural Values, Memory, and Nostalgia amidst Suburban Development in the American South. Master’s Thesis. Department of Anthropology, Hunter College. Available online at http://academicworks.cuny.edu/cgi/viewcontent.cgi?article=1127&context=hc_sas_etds.

Ripple, WJ, et al. (2015) Collapse of the world’s largest herbivores. Science Advances, 1(4):e1400103.

Rocha J, J Yletyinen, R Biggs, T Blenckner, and G Peterson (2015) Marine regime shifts: drivers and impacts on ecosystems services. Phil. Trans. R. Soc. B, 370: 20130273. http://dx.doi.org/10.1098/rstb.2013.0273

Rockstrom J, et al. (2009a) Planetary boundaries: exploring the safe operating space for humanity. Ecology and Society, 14(2): 32. http://www.ecologyandsociety.org/vol14/iss2/art32/

Rockstrom J, et al. (2009b) A safe operating space for humanity. Nature, 461:472–475.

Rohlf FJ (1998) On applications of geometric morphometrics to studies of ontogeny and phylogeny. Systematic Biology, 47:147–158.

Rohlf FJ (2003) Bias and error in estimates of mean shape in geometric morphometrics. Journal of Human Evolution, 44(6):665–683.

Rotz S and EDG Frase (2015) Resilience and the industrial food system: analyzing the impacts of agricultural industrialization on food system vulnerability. J Environ Stud Sci, 5:459–473.

Rozins C and T Day (2017) The industrialization of farming may be driving virulence evolution. Evolutionary Applications, 10(2):189–198.

Sauer S (2018) Soy expansion into the agricultural frontiers of the Brazilian Amazon: The agribusiness economy and its social and environmental conflicts. Land Use Policy, 79:326–338.

Schizas D (2012) Systems ecology reloaded: A critical assessment focusing on the relations between science and ideology. In G.P. Stamou (ed), Populations, Biocommunities, Ecosystems: A Review of Controversies in Ecological Thinking. Bentham Science Publishers, Sharjah.

Schlager S (2017) Morpho and Rvcg - Shape analysis in R. In G Zheng, S Li, and G Szekely (eds), Statistical Shape and Deformation Analysis. Academic Press, pp 217–256.

Schneider M (2017) Wasting the rural: Meat, manure, and the politics of agro-industrialization in contemporary China. Geoforum, 78:89–97.

Sharp G (2016) Metabolic rift theory and the crisis of our foodways. In JS Ormrod (ed), Changing Our Environment, Changing Ourselves: Nature, Labour, Knowledge and Alienation. Springer, Cham.

Shill HB (1992) Information “publics” and equitable access to electronic government information: The case of agriculture. Government Information Quarterly, 9(3):305–322.

Smith N (2008) Uneven Development: Nature, Capital, and the Production of Space. Third Edition. University of Georgia Press, Athens, GA.

Song X-P, C Huang, SS Saatchi, MC Hansen, and JR Townshend (2015) Annual carbon emissions from deforestation in the Amazon Basin between 2000 and 2010. PLoS ONE, 10(5): e0126754.

Stehle S and R Schulz (2015) Agricultural insecticides threaten surface waters at the global scale. PNAS, 112(18):5750–5755.

Tessum CW, JS Apte, A. Goodkind, NZ Muller, KA Mullins, et al. (2019) Inequity in consumption of goods and services adds to racial–ethnic disparities in air pollution exposure. PNAS, 116(13):6001–6006.

Tobler W (1994) Bidimensional regression. Geographical Analysis, 26(3):187–211.

Tumber C (2019) Land without bread: The Green New Deal forsakes America’s countryside. The Baffler. https://thebaffler.com/salvos/land-without-bread-tumber.

USDA-Economic Research Service (2019) Assets, debt, and wealth. https://www.ers.usda.gov/topics/farm-economy/farm-sector-income-finances/assets-debt-and-wealth/.

USDA-Economic Research Service (2019) Farming and farm income.https://www.ers.usda.gov/data-products/ag-and-food-statistics-charting-the-essentials/farming-and-farm-income/

USDA-Farm Service Agency (2014) Conservation Reserve Program, Summary and Enrollment Statistics, Fiscal Year 2012. https://www.fsa.usda.gov/Assets/USDA-FSA-Public/usdafiles/Conservation/PDF/crpstat0912.pdf.

USDA-National Agricultural Statistics Service (2012) Agricultural Statistics 2012. https://www.nass.usda.gov/Publications/Ag_Statistics/2012/2012_Ag_Stat.pdf

USDA-National Agricultural Statistics Service (2014a) 2012 Census of Agriculture. United States Summary and State Data Volume 1. Geographic Area Series. Part 51AC-12-A-51.

USDA-National Agricultural Statistics Service (2014b) Crop Production, 2013 Summary.

Vandermeer J, A Aga, J Allgeier, C Badgley, R Baucom, J Blesh, LF Shapiro, AD Jones, L Hoey, M Jain, I Perfecto, and ML Wilson (2018) Feeding Prometheus: An interdisciplinary approach for solving the global food crisis. Front. Sustain. Food Syst., 23 July, https://doi.org/10.3389/fsufs.2018.00039

Van Dijk G, JT van Dissel, P Speelman, JA Stegeman, P Vanthemsche, J de Vries, and CMJ van Woerkum (2010) Van Verwerping tot Verheffing: Q-koortsbeleid in Nederland 2005-2010. Evaluatiecommissie Q-koorts. https://www.burgemeesters.nl/ffles/File/Crisisbeheersing/docs/20101122.pdf.

Vesterby M and KS Krupa (1997) Major Uses of Land in the United States, 1997. Resource Economics Division, Economic Research Service,U.S. Department of Agriculture. Statistical Bulletin No. 973.

Wallace R, L Bergmann, L Hogerwerf, R Kock, and RG Wallace (2016) Ebola in the hog sector: Modeling pandemic emergence in commodity livestock. In RG Wallace and R Wallace (eds), Neoliberal Ebola: Modeling Disease Emergence from Finance to Forest and Farm. Springer, Cham.

Wallace R, LF Chaves, LR Bergmann, C Ayres, L Hogerwerf, R Kock, and RG Wallace (2018) Clear-Cutting Disease Control: Capital-Led Deforestation, Public Health Austerity, and Vector-Borne Infection. Springer, Cham.

Wallace R, A Liebman, L Bergmann, and RG Wallace. Agribusiness vs. public health: Disease control in resource-asymmetric conflict. Social Science & Medicine. Submitted.

Wallace RG (2002) The shape of space: applying geometric morphometrics to geographic data. Environment and Planning A, 34:119–144.

Wallace RG (2009) Breeding influenza: the political virology of offshore farming. Antipode, 41:916–951.

Wallace RG (2011) Occupy mathematics. Farming Pathogens blog, November 8. https://farmingpathogens.wordpress.com/2011/11/08/occupy-mathematics/.

Wallace RG (2016a) Pale, mushy wing. In Big Farms Make Big Flu: Dispatches on Infectious Disease, Agribusiness, and the Nature of Science. Monthly Review Press, New York, pp 222–223.

Wallace RG (2016b) Protecting H3N2v’s privacy. In Big Farms Make Big Flu: Dispatches on Infectious Disease, Agribusiness, and the Nature of Science. Monthly Review Press, New York, pp 319–321.

Wallace RG (2018a) Vladimir Iowa Lenin 1: A Bolshevik’s study of American agriculture. Capitalism Nature Socialism, 29(2):92–107.

Wallace RG (2018b) Vladimir Iowa Lenin 2: On rural proletarianization and an alternate food future. Capitalism Nature Socialism, 29(3):21–35.

Wallace RG (2018c) Duck and Cover: Epidemiological and Economic Implications of Ill-founded Assertions that Pasture Poultry Are an Inherent Disease Risk. Australian Food Sovereignty Alliance. https://afsa.org.au/wpcontent/uploads/2018/10/WallaceDuck-and-CoverReport-September2018.pdf.

Wallace RG, R Alders, R Kock, T Jonas, R Wallace, and L Hogerwerf (2019) Health before medicine: Community resilience in food landscapes. In M Walton (ed) One Planet, One Health: Looking After Humans, Animals and the Environment. Sydney University Press, Sydney.

Wallace RG, L Bergmann, R Kock, M Gilbert, L Hogerwerf, R Wallace and M Holmberg (2015) The dawn of Structural One Health: A new science tracking disease emergence along circuits of capital. Social Science & Medicine, 129:68–77.

Wallace RG and RA Kock (2012) Whose food footprint? Capitalism, agriculture and the environment. Human Geography, 5(1): 63–83.

Wallace RG, A Liebman, D Weisberger, T Jonas, L Bergmann, R Kock, and R Wallace. Industrial agricultural environments. The Routledge Handbook of Biosecurity and Invasive Species. Routledge, New York. In preparation.

Wallace RG, K Okamoto, and A Liebman (In press) Earth, the alien planet. In DB Monk and M Sorkin (eds) Between Catastrophe and Redemption: Essays in Honor of Mike Davis. UR Books, New York.

Wallace-Wells D (2019) The Uninhabitable Earth: Life After Warming. Tim Duggan Books, New York.

Weber JG and N Key (2014) Do wealth gains from land appreciation cause farmers to expand acreage or buy land? American Journal of Agricultural Economics, 96(5):1334–1348.

Weber MB (2018) Manufacturing the American Way of Farming: Agriculture, Agribusiness, and Marketing in the Post-war Period. Doctoral dissertation, Department of Rural, Agricultural, Technological, and Environmental History, Iowa State University. Available online at https://lib.dr.iastate.edu/cgi/viewcontent.cgi?article=7492&context=etd.

Winther RG (2006) On the dangers of making scientific models ontologically independent: taking Richard Levins’ warnings seriously, Biol Philos, 21:703–724.

Wooten HH (1953) Major Uses of Land in the United States. TB-1082. Bureau of Agricultural Economics, U.S. Department of Agriculture.

Wooten HH and JR Anderson (1957) Major Uses of Land in the United States: Summary for 1954. AIB-168, Agricultural Research Service, U.S. Department of Agriculture.

Wooten HH, K Gertel, and WC Pendleton (1962) Major Uses of Land and Water in the United States: Summary of 1959. AER-13. Economic Research Service, U.S. Department of Agriculture.

Xuan L, D Baotong, and Y Hua (2011) The research based on the 3-R principle of agro-circular economy model: The Erhai Lake Basin as an example. Energy Procedia, 5:1399–1404.

Yabe N (2005) The analysis of isoline of land price in the Tokyo metropolitan area applying geometric morphometrics. Theory and Applications of GIS, 13(2):11–20.

Yuan ZW, B Jun, and YC Moriguichi (2006). The circular ecology: a new development strategy in China. Journal of Industrial Ecology, 10:4–8.

Zhou Z, C Wang, and Y Luo (2018) Effects of forest degradation on microbial communities and soil carbon cycling: A global meta-analysis. Global Ecology and Biogeography, 27(1):110–124.

